# Increased transmissibility of emerging SARS-CoV-2 variants is driven either by viral load or probability of infection rather than environmental stability

**DOI:** 10.1101/2021.07.19.21260707

**Authors:** Yehuda Arav, Eyal Fattal, Ziv Klausner

## Abstract

Understanding the factors that increase the transmissibility of the recently emerging variants of SARS-CoV-2 (such as the Alpha, Epsilon, and Delta variants) can aid in mitigating their spread. The enhanced transmissibility could be attributed to one or more factors: higher stability on surfaces or within droplet nuclei suspended in air, increased maximal viral load or higher probability of infection. The relative importance of these factors on the transmission was examined using a validated stochastic-jump-continuous hybrid model. The transmissibility was quantified in terms of the household secondary attack rate (hSAR) which is the probability of transmission from an infected individual to a susceptible one in a household. We find that an increase in either the maximal viral load or the probability of infection is consistent with the observed hSAR of the variants. Specifically, in order to reach the relative increase in the hSAR of 40%, 55%, and 87% reported for the Epsilon, Alpha, and Delta variants (respectively), the maximal viral load should increase by 56%, 78%, and 125%, respectively. Alternatively, the probability of infection should increase by 34%, 53%, and 193%, respectively. Contrary to these results, even a dramatic increase in environmental stability increases hSAR by no more than 10%.

## Introduction

Since December 2020 the genomic surveillance effort in many countries has led to the detection of numerous variants of SARS-Cov-2^1–3^, some of which have exhibited an increased transmissibility. These variants have raised concerns in the public health authorities worldwide due to the risk that they will spread faster than vaccine production and distribution^2, 4, 5^. Understanding the mechanism that enhances the transmissibility of these variants is an important step in devising methods to control their transmission^6^.

Respiratory viruses such as SARS-CoV-2 propagate via four modes of transmission^7^ (Figure 1): direct physical contact between people (Figure 1, mode 1); indirect physical contact via small, frequently touched objects (fomites), such as a doorknob or a faucet, (Figure 1, mode 2); touching environmental surfaces, such as furniture, contaminated by droplets (Figure 1, mode 3); inhalation of droplet nuclei suspended in the air (Figure 1, mode 4).

**Figure 1.**
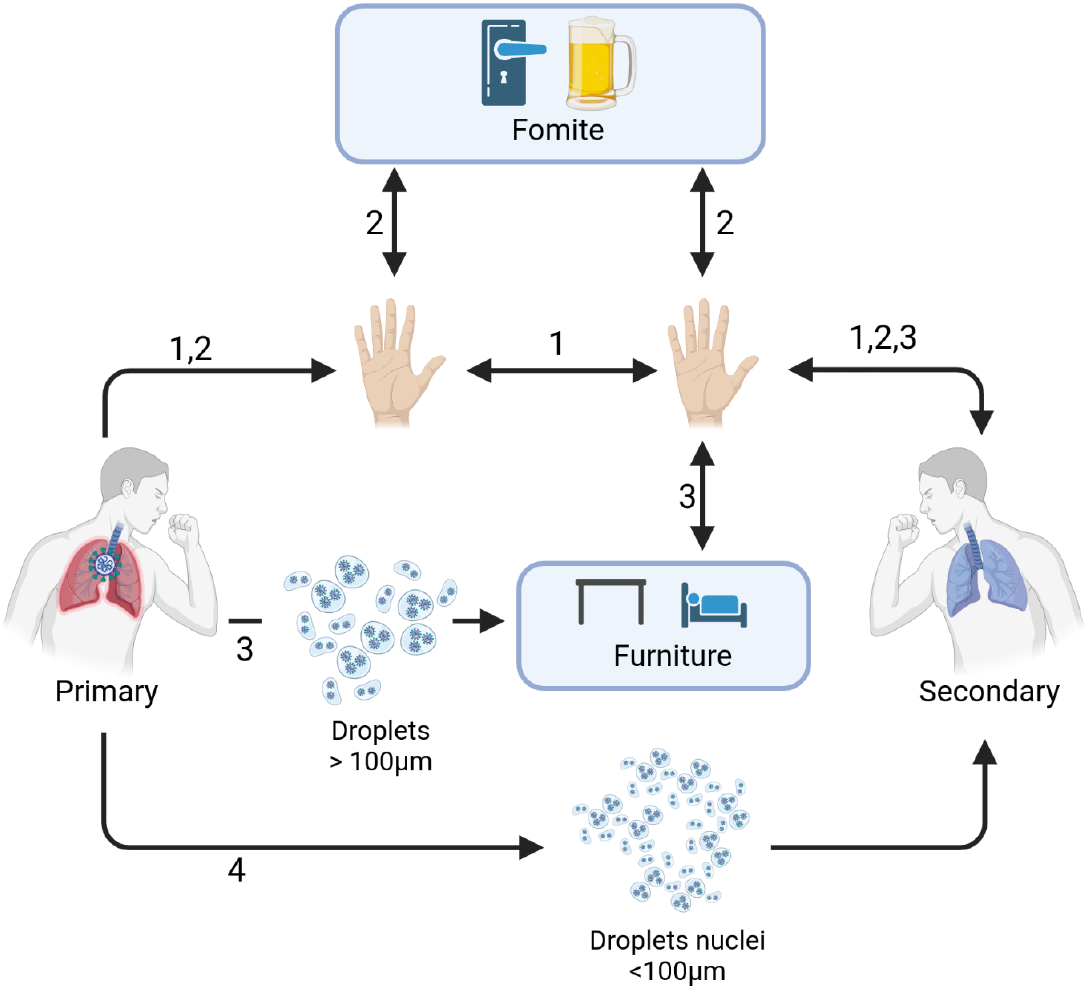
Schematic representation of the modes of transmission from the primary (infector) and secondary (infectee) individuals. (1) Direct contact (2) Indirect contact via fomites (3) Indirect contact via environmental surfaces (4) droplet nuclei (Created with BioRender.com).

Genetic variations that affect any of the modes of transmission might be the factor that increases the transmissibility of a SARS-CoV-2 variant compared to the wild-type, which is the previously dominating virus strain^6^. Specifically, increased stability on surfaces, would increase the number of virus copies that survive on contaminated surfaces (such as hands, fomites, or environmental surfaces) and would increase the transmission through the direct (transmission mode 1) and indirect modes (transmission modes 2 and 3)^8^, hereafter referred to as factor 1; increased stability in droplet nuclei would increase the transmission via mode 3^8^, hereafter referred to as factor 2; increased maximal viral load^4, 9^ would increase the viral shedding (the contagiousness of the primary) and thereby increase the transmission through all the modes^4, 9^, hereafter referred to as factor 3; probability of infection by a single plaque forming unit (PFU), meaning that a smaller number of virus copies is required to initiate an infection (increase the susceptibility of the secondary)^4^, hereafter referred to as factor 4. We note that the spread of variants whose increased transmissibility is due to factor 1 or 2 could be mitigated by adhering to stricter regimes of various hygienic and behavioral measures.

There are several ways to characterize the transmissibility of a new virus variant. It can be quantified as the effective reproduction number of the invading new variant and compared to that of the wild-type^10^. This comparison is often expressed in relative terms^9, 11^. Another possibility is to describe transmissibility by estimating the secondary attack rate (SAR) defined as the probability of an infected person to infect a susceptible person^12, 13^. Often, the SAR is stratified by the different environments in which people may be exposed to an infected person, e.g., public transportation, healthcare and households^14, 15^. Among these, it was found that household settings are associated with high risk of infection by the SARS-CoV-2 virus^14, 16, 17^. This can be illustrated by the fact that during the first wave of the COVID-19 pandemic in Israel, 65.8% of the cases were infected at home^18^. The household SAR (hSAR) is closely related to the reproduction number^17, 19^, several studies have characterised the increased transmissibility of a variant as the increase in hSAR relative to the wild-type. For example, the hSAR of the Alpha variant (lineage B.1.1.7, first detected in England^20^) was reported to increase by 55% in Oslo, Norway, and the hSAR of the Epsilon variant (lineages B.1.427 and B.1.429, “California” or “West Coast” variants^20, 21^) increased by 40% in San Francisco, United States^21, 22^. Recent information from the United Kingdom on the hSAR of the Delta variant (lineage B.2.627.2, first detected in India,^20^) leads to an estimated relative increase of 87% with respect to the wild-type that preceded the Alpha variant (_23, 24_).

The aim of this work is to identify the factor (or factors) that can lead to the higher transmissibility of an emerging SARS-CoV-2 variant.

## Methods

### Mathematical model

A mechanistic mathematical model that describes SARS-CoV-2 transmission in a household^25^ was used in order to examine the effect of each of the four factors on the transmission of SARS-CoV-2 variant. The model tracks the transfer of SARS-CoV-2 copies from a primary individual to a secondary individual via four modes of transmission (direct contact, indirect contact, droplet and droplet nuclei, Figure 1). An outline of the model is presented here. The detailed model equations, parameters and validation are described in Arav, et al.^25^.

The model equations are formulated in a hybrid continuous and stochastic-jump framework^26^ in order to take into account two distinct dynamical regimes: fast-discrete random events that represent the actions of the individuals (such as coughing, talking, touching), and slow-continuous events (such as the decay of the virus on surfaces, hands, and in the air). In this framework, the actions of the individuals are described as stochastic jump Poisson processes, and represent behavior patterns that typically occur in the living room, kitchen, bath, and bedrooms. The model describes the transfer of virus copies that result from each action, and consequently it is not necessary to follow the specific location of each individual. The environmental decay processes of the virus on the hands and on surfaces are described using continuum dynamics.

Since the actions of the individuals are represented as a stochastic process, we conducted a Monte Carlo simulation in which multiple realizations were computed to obtain the appropriate ensemble statistics. Thus it is possible to explicitly calculate higher order statistics crucial for the reconstruction of the observed serial interval distribution as well as the time dependant probability of infection^25^. The model validation was performed on data prior to December 2020, so it can be considered as describing the wild-type strain, before the emergence of the highly transmissible lineages.

### Modeling the effect of the factors on the transmissibility

The examination of the effect of genetic variations that increase the stability of the variant in the environment was divided into two parts: genetic variations that increase the stability on surfaces (factor 1) and in the droplet nuclei that are suspended in the air (factor 2). The effect of factor 1 was examined by decreasing the decay rate on furniture (*α*_*furniture*_), fomites (*α*_*fomite*_) and on the hands of the primary and the secondary individuals (*α*_*hand*_). The effect of factor 2 was examined by decreasing the decay rate of the virus in the aerosol (*α*_*air*_). The benchmark values that represent the wild-type were *α*_*air*_, *α*_*furniture*_, and *α*_*fomite*_ of 1, 6, and 6 1/h, respectively^27^. In order to quantify the effect of either factor 1 or 2 relative to the wild-type, the relevant decay rates were divided by values, from 2 up to 8. It should be noted that *α*_*furniture*_ and *α*_*fomite*_ were set to equal values, based on the fact that the difference between environmental and fomite surfaces lies in their area and the rate that each type of surface is touched, but not in the decay rate of virus copies that are deposited on them^25^. In order to examine the effect of factor 3, the maximal viral load (*L*_*max*_), we conducted simulations with a relative increase of this parameter, from 10% up to 100% with respect to the wild-type. The benchmark value of *L*_*max*_ was 2 · 10^8^ copies^28^. The effect of factor 4, genetic variations that increase the probability of infection by a single PFU was examined by increasing the reciprocal of the dose-response coefficient *k*. For the SARS-CoV-2 wild-type, the value of *k* was 410 PFU, which translates to 1.2 · 10^5^ viral copies^29, 30^. The effect of factor 4 was examined by performing model simulation with a relative decrease in the parameter *k*, from 10% to 200%.

## Results

The increase of virus stability exhibited similar behavior regarding both stability on surfaces, factor 1, and in droplets nuclei suspended in the air, factor 2 (Figure 2). Relative decrease in the decay rates of up to 2 leads to a relative increase of the hSAR but only up to 10%. However, a further decrease of the decay rates does not lead to further increase of the hSAR. The relative increase of the hSAR of 10% is far even from the reported relative hSAR increase of 40% associated the Epsilon variant, let alone from the even more transmissible variants, Alpha and Delta^21–24^.

**Figure 2.**
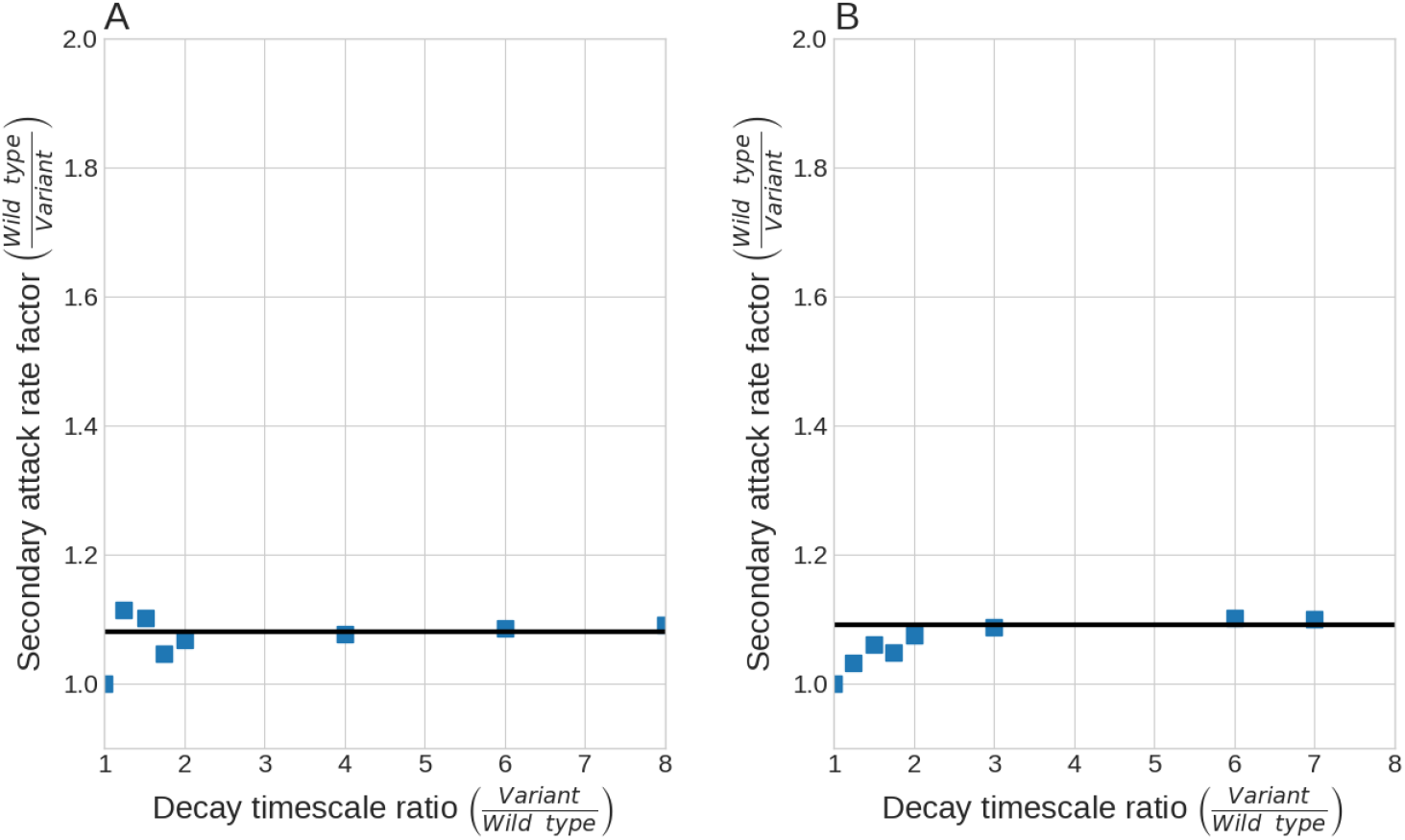
The effect of genetic variant with higher environment stability on the hSAR. (A) Factor 1 -The decay rates on surfaces (*α*_*furniture*_, *α*_*fomite*_, and *α*_*hand*_) on the hSAR ; (B) Factor 2 -The decay rate in the air (*α*_*air*_)

Regarding the effect of the maximal viral load (factor 3) and the probability of infection by a single PFU (factor 4), it was found that the relative increase in the relative parameters is linearly associated with the relative increase of the hSAR (Figure 3). In both cases, the association is a strong positive relationship (*R*^2^ = 0.99). Specifically, the fitted linear model describing the effect of factor 3 yields a slope of 0.68 and an intercept of 0.342 (*R*^2^ = 0.99), whereas the coefficients of the fitted regression model describing the effect of factor 4 are a slope of 0.79 and an intercept of 0.342 (*R*^2^ = 0.99). Using these linear relationships it is possible to estimate the relative increase in each of these factors in order to reach the relative increase in the hSAR of 40%, 55%, and 87% reported for the Epsilon, Alpha, and Delta variants, respectively^21–24^. If the increased transmissibility of the Epsilon, Alpha and Delta is driven by increase in factor 3, the maximal viral load should increase by 56%, 78%, and 125%, respectively. Regarding factor 4, the probability of infection by a single PFU should increase by 34%, 53%, and 193%, respectively.

**Figure 3.**
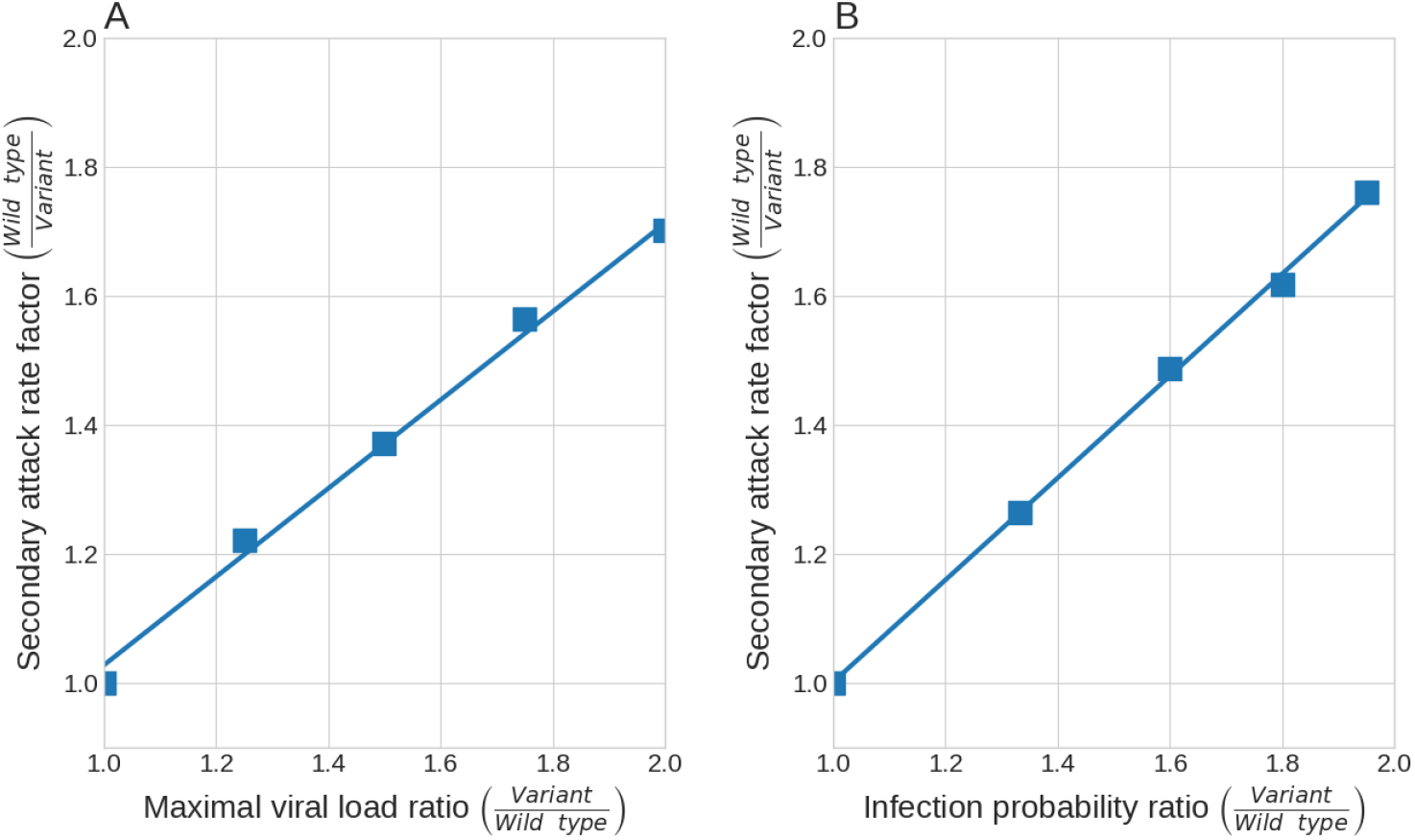
(A) The effect of the maximal viral load on the SAR. (B) The effect of infection probability on the SAR.

It should be noted that the increase of the hSAR resulted by the increase of either factors 3 and 4, does not change the relative contribution of each mode to the overall transmission. This is similar to the relative contribution in the wild-type, that is, approximately 70% of the copies transfer by direct contact, 20% by indirect contact of fomites, 10% by the droplet nuclei mode, and transmission by environmental surface is negligible^25^.

## Discussion and conclusions

Analysis of the model results showed that variants with enhanced environmental stability (either on surfaces, factor 1, or in the air, factor 2) exhibit only a modest increase of up to 10% in the hSAR (Figure 2A and B, respectively). Since the hSAR of the Alpha, Epsilon, and Delta variants is 40% *−* 87% higher than the wild-type^20–22,24^, we conclude that increased environmental stability could not be the factor that drives the increased transmissibility of these variant. This conclusion is consistent with the results of Schuit, et al.^8^, that found that the stability of SARS-CoV-2 aerosols does not vary greatly among circulating lineages, including the Alpha variant, thus indicating that the increased transmissibility is not due to enhanced environmental survival. The results regarding factor 1 can be explained due to the fact that the survival of virus on the surfaces is not the rate limiting step in the direct and indirect transfer. Indeed, Arav, et al.^25^ have shown that the rate limiting step in direct and indirect contact is the frequency of contact events on fomites and the face, which has a period of approximately 30 to 40 minutes. Regarding factor 2, can be attributed to the fact that the concentration of the droplet nuclei in the air is determined by the ventilation rate of the household which is 0.3*h*^*−*1^ ^31, 32^

Alternatively to the limited effect of factors 1 and 2, we have found that the hSAR increases linearly (coefficient of 0.7 *−* 0.8) with the maximal viral load, factor 3, and the probability of infection by a single PFU, factor 4 (Figure 3A and B, respectively). Moreover, we were able to estimate the relative change in each factor that may lead to the observed increase of the hSAR of the Epsilon, Alpha, and Delta variants. These results support the possibility that genetic variations that increase either the contagiousness of the primary individual (factor 3) or the susceptibility of the secondary individual (factor 4) may be responsible for the observed increase in the transmissibility of these three variants. While these two factors have comparable impact on the hSAR, it should be noted regarding the Alpha and Epsilon variants, it was reported that the viral load associated with these variants was not significantly different from the viral load of other circulating strains^21, 33^. As these reports rule out the possibility of factor 3, it leaves factor 4 as the only factor that can lead to the increased transmissibility of the Epsilon and Alpha variants. While the exact mechanism by which genetic variations increase the probability of infection by a single PFU is not currently known, Nelson, et al.^34^ and Gan, et al.^35^ have shown that certain variations in the spike receptor binding domain (S RBD) increase the affinity to the human angiotensin-converting enzyme 2 (hACE2) and thereby promote the entry to the cell. This enhanced affinity of the S RBD to the hACE2 is likely to explain the greater transmissibility of many variants. It seems reasonable that the manifestation of this affinity is the increased probability of infection by a single PFU.

This study found that the increase in the transmissibility is not due to environmental stability. Therefore, from a public health point of view, leading even stricter hygienic and behavioral measures are not expected to achieve a pronounced mitigating effect. However, the results that factors that possibly drive the increased transmissibility were found to be thos that affect all modes of transmission, may strengthen the importance of wearing masks in indoors environments.

## Data Availability

All relevant data is included in the manuscript

## Author contributions statement

YA conceived the study, performed the formal analysis, developed the methodology, software and visualization, wrote the original draft and critically revised the manuscript. EF conceived the study, developed the methodology, acquired the funding and the computational infrastructure, and critically revised the manuscript. ZK conceived the study, performed the formal analysis, developed the methodology, wrote the original draft and critically revised the manuscript. All authors gave final approval for publication and agree to be held accountable for the work performed therein.

## References

1. Lauring, A. S. & Hodcroft, E. B. Genetic Variants of SARS-CoV-2 - What Do They Mean? JAMA - J. Am. Med. Assoc. 325, 529–531, DOI: 10.1001/jama.2020.27124 (2021).

2. Walensky, R. P., Walke, H. T. & Fauci, A. S. SARS-CoV-2 Variants of Concern in the United States-Challenges and Opportunities. JAMA - J. Am. Med. Assoc. 325, 1037–1038, DOI: 10.1001/jama.2021.2294 (2021).

3. Moelling, K. Within-Host and Between-Host Evolution in SARS-CoV-2—New Variant’s Source. Viruses 13, 751, DOI: 10.3390/v13050751 (2021).

4. Grubaugh, N. D., Hodcroft, E. B., Fauver, J. R., Phelan, A. L. & Cevik, M. Public health actions to control new SARS-CoV-2 variants. Cell 184, 1127–1132, DOI: 10.1016/j.cell.2021.01.044 (2021).

5. Gonzalez-Parra, G., Martínez-Rodríguez, D. & Villanueva-Micó, R. Impact of a New SARS-CoV-2 Variant on the Population: A Mathematical Modeling Approach. Math. Comput. Appl. 26, 25, DOI: 10.3390/mca26020025 (2021).

6. Leung, N. H. Transmissibility and transmission of respiratory viruses, DOI: 10.1038/s41579-021-00535-6 (2021).

7. Kutter, J. S., Spronken, M. I., Fraaij, P. L., Fouchier, R. A. & Herfst, S. Transmission routes of respiratory viruses among humans, DOI: 10.1016/j.coviro.2018.01.001 (2018).

8. Schuit, M. et al. The stability of an isolate of the SARS-CoV-2 B.1.1.7 lineage in aerosols is similar to three earlier isolates. The J. Infect. Dis. 1–18, DOI: 10.1093/infdis/jiab171 (2021).

9. Davies, N. G. et al. Estimated transmissibility and impact of SARS-CoV-2 lineage B.1.1.7 in England. Science 372, eabg3055, DOI: 10.1126/science.abg3055 (2021).

10. Volz, E. et al. Assessing transmissibility of SARS-CoV-2 lineage B.1.1.7 in England. Nature 593, 266–269, DOI: 10.1038/s41586-021-03470-x (2021).

11. Munitz, A., Yechezkel, M., Dickstein, Y., Yamin, D. & Gerlic, M. BNT162b2 vaccination effectively prevents the rapid rise of SARS-CoV-2 variant B.1.1.7 in high-risk populations in Israel. Cell Reports Medicine 100264, DOI: 10.1016/j.xcrm.2021.100264 (2021).

12. Park, J. E. & Ryu, Y. Transmissibility and severity of influenza virus by subtype. Infect. Genet. Evol. 65, 288–292, DOI: 10.1016/j.meegid.2018.08.007 (2018).

13. Black, A. J., Geard, N., McCaw, J. M., McVernon, J. & Ross, J. V. Characterising pandemic severity and transmissibility from data collected during first few hundred studies. Epidemics 19, 61–73, DOI: 10.1016/j.epidem.2017.01.004 (2017).

14. Luo, L. et al. Modes of contact and risk of transmission in COVID-19 among close contacts. Digit. Econ. at Glob. Margins 2020.03.24.20042606, DOI: 10.7551/mitpress/10890.003.0026 (2020).

15. Koh, W. C. et al. What do we know about SARS-CoV-2 transmission? A systematic review and meta-analysis of the secondary attack rate and associated risk factors. PLoS ONE 15, 1–23, DOI: 10.1371/journal.pone.0240205 (2020).

16. Bi, Q. et al. Epidemiology and Transmission of COVID-19 in Shenzhen China: Analysis of 391 cases and 1,286 of their close contacts. medRxiv 2020.03.03.20028423, DOI: 10.1101/2020.03.03.20028423 (2020).

17. Madewell, Z. J., Yang, Y., Longini, I. M., Halloran, M. E. & Dean, N. E. Household Transmission of SARS-CoV-2: A Systematic Review and Meta-analysis. JAMA network open 3, e2031756, DOI: 10.1001/jamanetworkopen.2020.31756 (2020).

18. Koch-Davidovich, F. The Israeli Ministry of Health data on the source of exposure of COVID-19 patients to the virus. Tech. Rep., Knesset, Jerusalem (2020).

19. Yang, S., Lee, G. W., Chen, C. M., Wu, C. C. & Yu, K. P. The size and concentration of droplets generated by coughing in human subjects. J. Aerosol Medicine: Deposition, Clear. Eff. Lung 20, 484–494, DOI: 10.1089/jam.2007.0610 (2007).

20. SARS-CoV-2 variants of concern and variants under investigation in England - Technical briefing 14. Tech. Rep. (2021).

21. Peng, J. et al. Estimation of secondary household attack rates for emergent spike L452R SARS-CoV-2 variants detected by genomic surveillance at a community-based testing site in San Francisco. Clin. Infect. Dis. DOI: 10.1093/cid/ciab283 (2021).

22. Lindstrøm, J. C. et al. Increased transmissibility of the B. 1. 1. 7 SARS-CoV-2 variant : Evidence from contact tracing data in Oslo, January to February 2021. medRxiv DOI: 10.1101/2021.03.29.21254122 (2021).

23. Investigation of novel SARS-CoV-2 variant Variant of Concern 202012/01 Technical briefing 3. 2021.

24. SARS-CoV-2 variants of concern and variants under investigation in England - Technical briefing 16. Tech. Rep. (2021).

25. Arav, Y., Klausner, Z. & Fattal, E. Theoretical investigation of pre-symptomatic SARS-CoV-2 person-to-person transmission in households. Sci. Reports 2021 11:1 11, 1–12, DOI: 10.1101/2020.05.12.20099085 (2021).

26. Tankov, P. Financial Modelling with Jump Processes. Financial Model. with Jump. Process. DOI: 10.1201/9780203485217 (2003).

27. van Doremalen, N. et al. Aerosol and Surface Stability of SARS-CoV-2 as Compared with SARS-CoV-1. New Engl. J. Medicine DOI: 10.1056/nejmc2004973 (2020).

28. Kleiboeker, S. et al. SARS-CoV-2 viral load assessment in respiratory samples. J. Clin. Virol. 129, 104439, DOI: 10.1016/j.jcv.2020.104439 (2020).

29. Watanabe, T., Bartrand, T. A., Weir, M. H., Omura, T. & Haas, C. N. Development of a dose-response model for SARS coronavirus. Risk Analysis 30, 1129–1138, DOI: 10.1111/j.1539-6924.2010.01427.x (2010).

30. Zhang, X. & Wang, J. Dose-response Relation Deduced for Coronaviruses From Coronavirus Disease 2019, Severe Acute Respiratory Syndrome, and Middle East Respiratory Syndrome: Meta-analysis Results and its Application for Infection Risk Assessment of Aerosol Transmission. Clin. Infect. Dis. 1–5, DOI: 10.1093/cid/ciaa1675 (2020).

31. Hou, J. et al. Air Change Rates in Residential Buildings in Tianjin, China. In Procedia Engineering, vol. 205, 2254–2258, DOI: 10.1016/j.proeng.2017.10.069 (Elsevier Ltd, 2017).

32. Yamamoto, N., Shendell, D. G., Winer, A. M. & Zhang, J. Residential air exchange rates in three major US metropolitan areas: Results from the Relationship among Indoor, Outdoor, and Personal Air Study 1999-2001. Indoor Air 20, 85–90, DOI: 10.1111/j.1600-0668.2009.00622.x (2010).

33. Walker, A. S. et al. Increased infections, but not viral burden, with a new SARS-CoV-2 variant. medRxiv 1–13, DOI: 10.1101/2021.01.13.21249721 (2021).

34. Nelson, G. et al. Molecular dynamic simulation reveals E484K mutation enhances spike RBD-ACE2 affinity and the 1 combination of E484K, K417N and N501Y mutations (501Y.V2 variant) induces conformational change greater than N501Y mutant alone, potentially resulting in an esc. bioRxiv DOI: 2021.01.13.426558 (2021).

35. Gan, H. H., Twaddle, A., Marchand, B. & Gunsalus, K. C. Structural Modeling of the SARS-CoV-2 Spike/Human ACE2 Complex Interface can Identify High-Affinity Variants Associated with Increased Transmissibility. J. Mol. Biol. 433, 167051, DOI: 10.1016/j.jmb.2021.167051 (2021).

